# Alternations in gut microbiota and host transcriptome of patients with coronary artery disease

**DOI:** 10.1101/2023.07.14.23292642

**Authors:** Liuying Chen, Xuanting Mou, Jingjing Li, Miaofu Li, Caijie Ye, Xiaofei Gao, Xiaohua Liu, Yunlong Ma, Yizhou Xu, Yigang Zhong

## Abstract

**Background:** Coronary artery disease (CAD) is a widespread heart condition caused by atherosclerosis and influences millions of people worldwide. Early detection of CAD is challenging due to the lack of specific biomarkers. The gut microbiota and host-microbiota interactions have been well documented to affect human health. However, investigation that reveals the role of gut microbes in CAD is still limited. This study aims to uncover the synergistic effects of host genes and gut microbes associated with CAD through integrative genomic analyses.

**Results:** Herein, we collected 54 fecal and 54 blood samples from CAD patients and matched controls, and performed amplicon and transcriptomic sequencing on these samples, respectively. By comparing CAD patients with health controls, we found that dysregulated gut microbes were significantly associated with CAD. By leveraging the Random Forest method, we found that 10 bacteria biomarkers can distinguish CAD patients from health controls with a high performance (AUC = 0.939). We observed that there existed prominent associations of gut microbes with several clinical indices relevant to heart functions. Integration analysis revealed that CAD-relevant gut microbe *genus Fusicatenibacter* was associated with expression of CAD-risk genes, such as *GBP2*, *MLKL*, and *CPR65*. In addition, the upregulation of immune-related pathways in CAD patients were identified to be primarily associated with higher abundance of genus *Blautia*, *Eubacterium*, *Fusicatenibacter*, and *Monoglobus*.

**Conclusions:** Our results highlight that dysregulated gut microbes contribute risk to CAD by interacting with host genes. These identified microbes and interacted risk genes may have high potentials as biomarkers for CAD.

## Background

Coronary artery disease (CAD) mostly arising from atherosclerosis is the most common type of heart disease, which affects millions of individuals worldwide [1]. The cause of CAD is multifactorial that involve complex environmental and genetic factors [2–4]. CAD process can be effectively prevented by drug therapy, percutaneous coronary intervention (PCI), or coronary artery bypass graft (CABG) surgery [5, 6]. However, the early diagnosis of CAD remains substantially difficult due to lack of inadequate biomarkers. Although several biomarkers, such as Fibrinogen and C-reactive protein, have been reported to be associated with CAD [7–10], these biomarkers generally lack sufficient specificity and significantly detectable changes appear mainly in the advanced stages of CAD [11]. Thus, the early diagnosis and intervention of CAD pose a significant public health challenge with enormous medical and societal consequences. There is an urgent requirement for new and effective biomarkers to assist in the early diagnosis, monitoring, and management of CAD.

Gut microbiota comprises the tens of thousands of intestinal bacteria and the biological activity in the human intestine [12–15], which is essential to the development of human health [16–24]. Host-microbiota interactions, including inflammatory and metabolic processes, have been well-documented to be involved in the etiology of multiple complex diseases [25, 26]. Multiple lines of evidence have shown that the gut microbiota can influence distant cells and organs via a variety of biochemical signals or metabolites [27]. In particular, recent studies have indicated that gut microbiome potentially impact the cardiovascular system [28, 29]. An increased potential for several intestinal flora metabolites, including lipopolysaccharides [30], grammabutyrobetaine [31], and trimethyllysine [32] biosynthesis in the microbiome, has been identified among CAD patients. Most notably, a growing number of studies have linked increased levels of the gut microbe-derived trimethylamine-N-oxide (TMAO) to cardiovascular diseases [33, 34]. It has also recently been demonstrated that the composition of gut microbiota can induce the alterations in various serum metabolite concentrations that are significantly relevant to the severity of CAD [35]. Hence, integrating metagenomics with other omics approaches including transcriptomics to investigate the host-microbiome interactions will allow for distinguishing effective biomarkers and developing precision medicine for the treatment of atherosclerotic cardiovascular diseases [36, 37]. To date, investigating the contribution of gut microbiome changes to CAD is insufficient, and the mechanism underlying synergistic interactions between host genes and gastrointestinal microbes remains unclear. Thus, we performed an integrative genomic analysis to uncover the synergy between host genes and gut microbes associated with CAD. Furthermore, we also conducted a series of bioinformatics analyses to explore the molecular functions and potential mechanisms of microbe-associated host genes for the development of CAD, and give a clue of the potential biomarkers for diagnosing CAD.

## Results

### Overview of gut microbiota

A total of 54 fecal and 54 blood samples comprising of 31 CAD patients and 23 controls were sequenced in the current study. Clinical information, including baseline characteristics, medications, and laboratory data of the CAD and control group, is shown in Table 1. There was no significant difference in age, gender, background medication, or other baseline characteristics between the two groups of subjects (P > 0.05).

**Table 1.** Clinical characteristics of patients with CAD and non-CAD.

|  | CAD<br>( n=31 ) | Control<br>( n=23 ) | *P* |
| --- | --- | --- | --- |
| Age (years) | 63.97±8.33 | 62.28±6.84 | 0.419 |
| Males (n, %) | 16 ( 51.61 ) | 12 ( 52.17 ) | 0.977 |
| Smoker (n, %) | 15 ( 48.39 ) | 9 ( 39.13 ) | 0.614 |
| Alcohol (n, %) | 14 ( 45.16 ) | 12 ( 52.17 ) | 0.713 |
| Arterial hypertension (n, %) | 18 ( 58.06 ) | 16 ( 69.57 ) | 0.598 |
| Diabetes mellitus (n, %) | 9 ( 29.03 ) | 5 ( 21.74 ) | 0.603 |
| Hyperlipidemia (n, %) | 4 ( 9.68 ) | 3 ( 13.04 ) | 0.989 |
| Family history of CAD (n, %) | 6 ( 19.35 ) | 2 ( 8.70 ) | 0.314 |
| Syncope (n, %) | 1 ( 3.23 ) | 0 ( 0 ) | 0.389 |
| Atrial fibrillation (n, %) | 2 ( 6.45 ) | 4 ( 17.39 ) | 0.233 |
| Peripheral arterial disease (n, %) | 1 ( 3.23 ) | 1 ( 4.35 ) | 0.832 |
| Leukocyte ( x10<sup>9</sup>/L ) | 6.26±1.62 | 6.07±1.91 | 0.782 |
| Neutrophil ( % ) | 63.14±7.51 | 61.82±9.81 | 0.571 |
| lymphocyte ( % ) | 27.58±6.57 | 29.08±8.06 | 0.445 |
| Hemoglobin ( g/L ) | 134.77±16.11 | 137.04±15.39 | 0.596 |
| Platelet ( x10<sup>9</sup>/L ) | 217.94±43.19 | 217.80±60.99 | 0.992 |
| Albumin ( g/L ) | 39.03±3.23 | 39.33±2.21 | 0.694 |
| ALT ( U/L ) | 22.81±11.30 | 18.25±5.31 | 0.074 |
| AST ( U/L ) | 23.55±7.27 | 23.63±4.45 | 0.962 |
| Creatinine ( umol/L ) | 76.74±18.39 | 73.03±19.73 | 0.475 |
| Uric acid ( $\mu\text{mol/L}$ ) | 310.35±85.91 | 305.35±81.14 | 0.829 |
| Triglyceride ( $\text{mmol/L}$ ) | 2.06±1.87 | 1.53±1.33 | 0.253 |
| Cholesterol ( $\text{mmol/L}$ ) | 4.13±0.87 | 4.21±0.93 | 0.736 |
| High-density lipoprotein<br>( $\text{mmol/L}$ ) | 1.05±0.25 | 1.17±0.27 | 0.099 |
| Low-density lipoprotein<br>( $\text{mmol/L}$ ) | 2.17±0.68 | 2.24±0.83 | 0.722 |
| CKMB ( $\text{U/L}$ ) | 14.26±9.46 | 14.83±4.91 | 0.788 |
| cTNI ( $\mu\text{g/L}$ ) | 0.05±0.14 | 0.01±0.00 | 0.168 |
| Echocardiography |  |  |  |
| LVEF ( % ) | 64.53±4.63 | 63.27±8.30 | 0.494 |
| Medication |  |  |  |
| ACEI (n, %) | 7 ( 22.58 ) | 3 ( 13.04 ) | 0.421 |
| ARB (n, %) | 10 ( 32.26 ) | 9 ( 39.13 ) | 0.674 |
| ARNI (n, %) | 3 ( 9.68 ) | 4 ( 17.39 ) | 0.436 |
| β-blocker (n, %) | 24 ( 77.42 ) | 12 ( 25.25 ) | 0.261 |
| Ivabradine (n, %) | 0 ( 0.00 ) | 2 ( 8.70 ) | 0.101 |
| MRA (n, %) | 7 ( 22.58 ) | 2 ( 8.70 ) | 0.217 |
| Antiplatelet<br>drugs/Anticoagulants (n, %) | 31 ( 100.00 ) | 23 ( 100.00 ) | 1.000 |
| Lipid lowering agents (n, %) | 28 ( 90.32 ) | 20 ( 86.96 ) | 0.897 |

Through 16S rRNA sequencing, we obtained 964 OTUs from all 54 fecal samples. Among them, there were 654 (67.84%) shared OTUs between CAD and control subjects, and 248 (25.73%) and 62 (6.43%) unique OTUs for CAD and control groups, respectively (Figure 1a). Although no significant difference was observed in Shannon (Figure 1b) and PD (Figure 1c) indexes between CAD and control groups, beta-diversity analysis revealed that there was a significant difference in overall bacterial composition between two groups (Figure 1d). To provide a clearer understanding of the gut microbial features in CAD patients, we summarized the relative abundance of gut microbes at different taxonomic levels. At phylum level, the major gut microbes belong to *Firmicutes*, *Bacteroidota*, *Fusobacteriota*, and *Proteobacteria* (Figure 1e and 1g). At genus level, the top 10 abundant bacteria were *Bacteroides*, *Prevotella_9*, *Lactobacillus*, *Limosilactobacillus*, *Fusobacterium*, *Ligilactobacillus*, *Escherichia-Shigella*, *Faecalibacterium*, *Klebsiella*, and *Alloprevotella* (Figure 1f).

**Figure 1.**
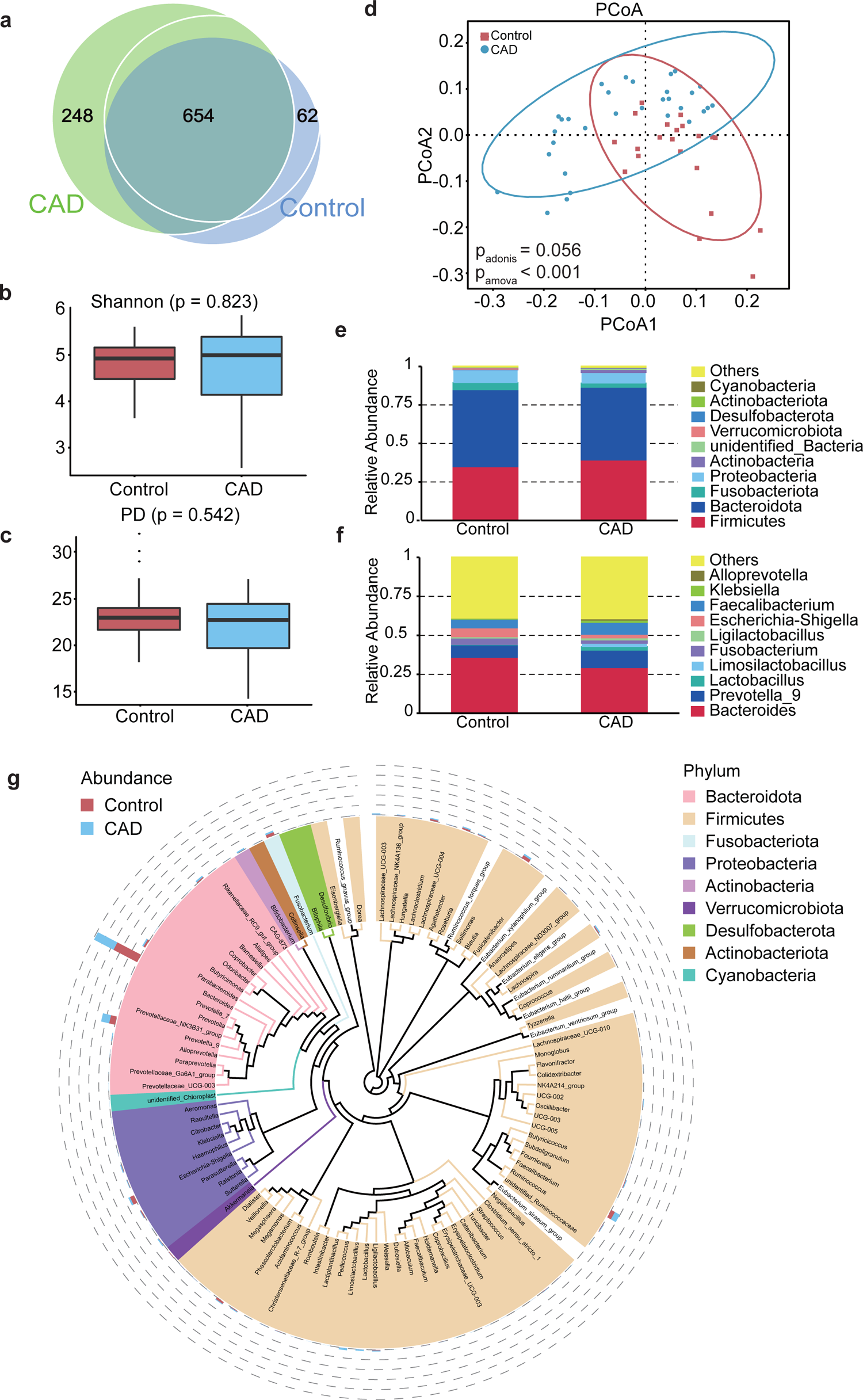
Overview of intestinal flora composition in control and CAD patients. (A) OTU Difference Venn diagram between Con and CAD; (B) and (C) The Shannon and PD indexes; (D) beta-diversity analysis; (E) and (F) the major gut microbes at phylum level; (G) the major gut microbes at genus level.

### Difference in gut microbiota between CAD and control group

By performing differential abundant analysis, we identified 18 differential abundant genera between CAD patients and health controls (Fig. 2a). Genus *Blautia*, *Fusicatenibacter*, *Monoglobus*, and *Eubacterium* were depleted in CAD patients, while *Sutterella*, *Lachnospiraceae_NK4A136_group*, *UCG-002*, *UCG-005*, *[Eubacterium]_hallii_grou*, *Collinsella*, *Colidextribacter*, *NK4A214_group*, *Negativibacillus*, *Faecalitalea*, *Family_XIII_AD3011_group*, *Peptoniphilus*, *Fructilactobacillus*, and *Solobacterium* were enriched in CAD patients. Among them, eight of these identified microbes, including *Blautia*, *Fusicatenibacter*, *Monoglobus*, *Eubacterium*, *UCG-002*, *UCG-005*, *Collinsella*, and *NK4A214_group*, have been reported to be associated with CAD, acute myocardial infarction, or CAD complicated with non-alcoholic fatty liver disease in previous studies [30, 38–45]. Notably, 10 microbes, including *Sutterella*, *Lachnospiraceae_NK4A136_group*, *[Eubacterium]_hallii_grou*, *Colidextribacter*, *Negativibacillus*, *Faecalitalea*, *Family_XIII_AD3011_group*, *Peptoniphilus*, *Fructilactobacillus*, and *Solobacterium*, were newly identified in the current investigation. Detailed taxonomic abundance differences of gut microbiota at all taxonomic levels are shown in the Supplementary Figure 1a-d.

**Figure 2.**
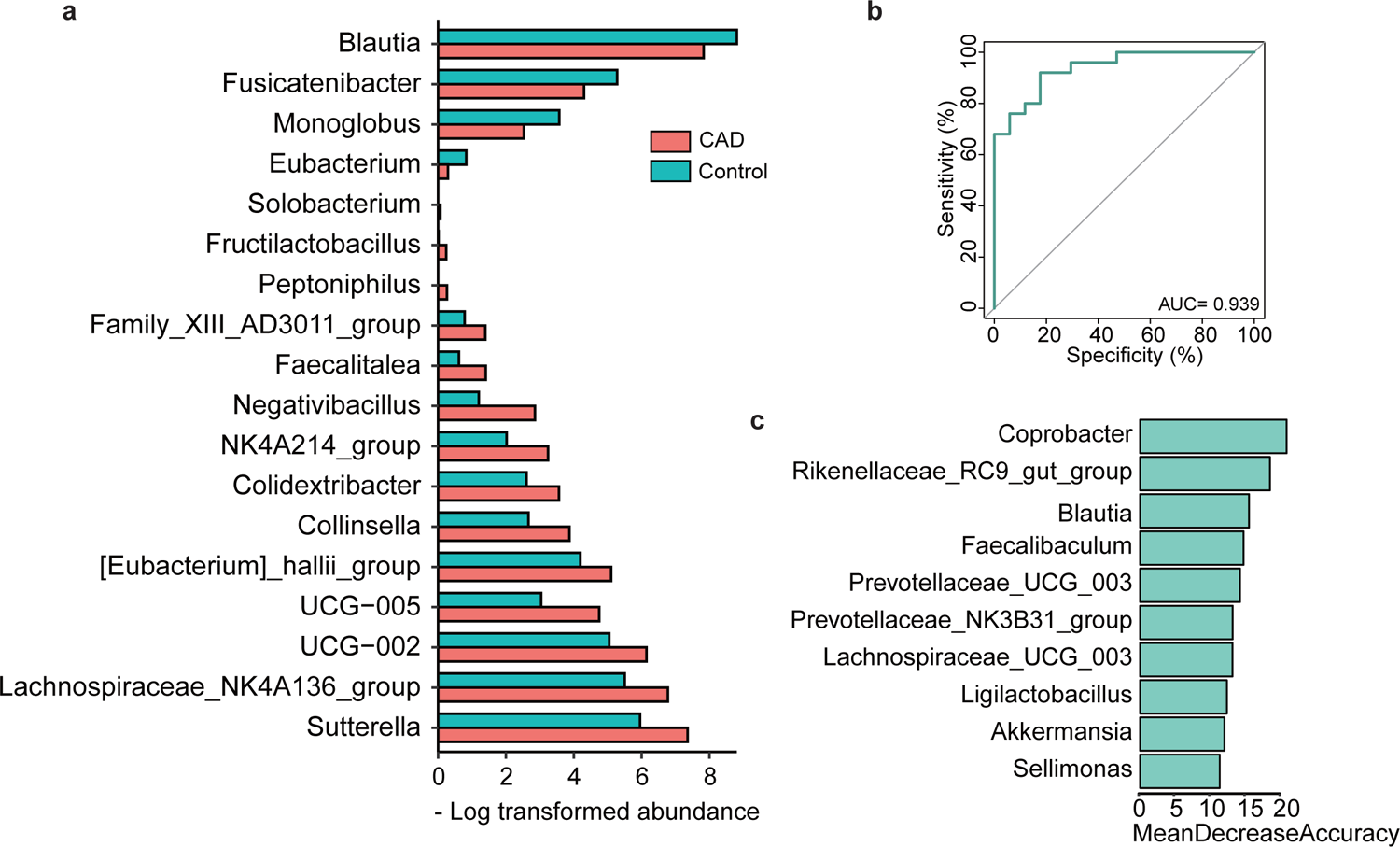
Difference of gut microbiota between CAD and Control group. (A) The differential abundant genera between CAD and control group; (B) The AUC of genus-level abundance. (C) The top 10 differential genera between CAD and control group.

By applying the Random Forest procedure, we found 10 bacteria biomarkers that can distinguish CAD patients from health controls. The overall AUC was 0.939 using genus-level abundance as input (Fig. 2b). By overlapping the top 10 genera identified from two independent methods of Mean Decrease Accuracy (Fig. 2c) and Mean Decrease Gini (Supplementary Fig. 1e), we emphasized the importance of *Coprobacter*, *Rikenellaceae_RC9_gut_group*, *Blautia*, and *Faecalibaculum* on CAD. Of note, the top-ranked biomarker of genus *Blautia* was also identified to be significantly differential genera.

### Functional implications of gut microbiota dysbiosis in CAD

To further explore gut microbial functional dysbiosis of CAD patients, we performed functional annotation and differential analyses based on bacterial profiles. We observed a depletion of pathways, including beta−Lactam resistance, cationic antimicrobial peptide (CAMP) resistance and viral proteins functions, and an up-regulation of pathways, including nitrogen andmethane metabolism, microRNAs in cancer and tropane, piperidine and pyridine alkaloid biosynthesis in CAD patients comparing with health controls (Fig. 3a).

**Figure 3.**
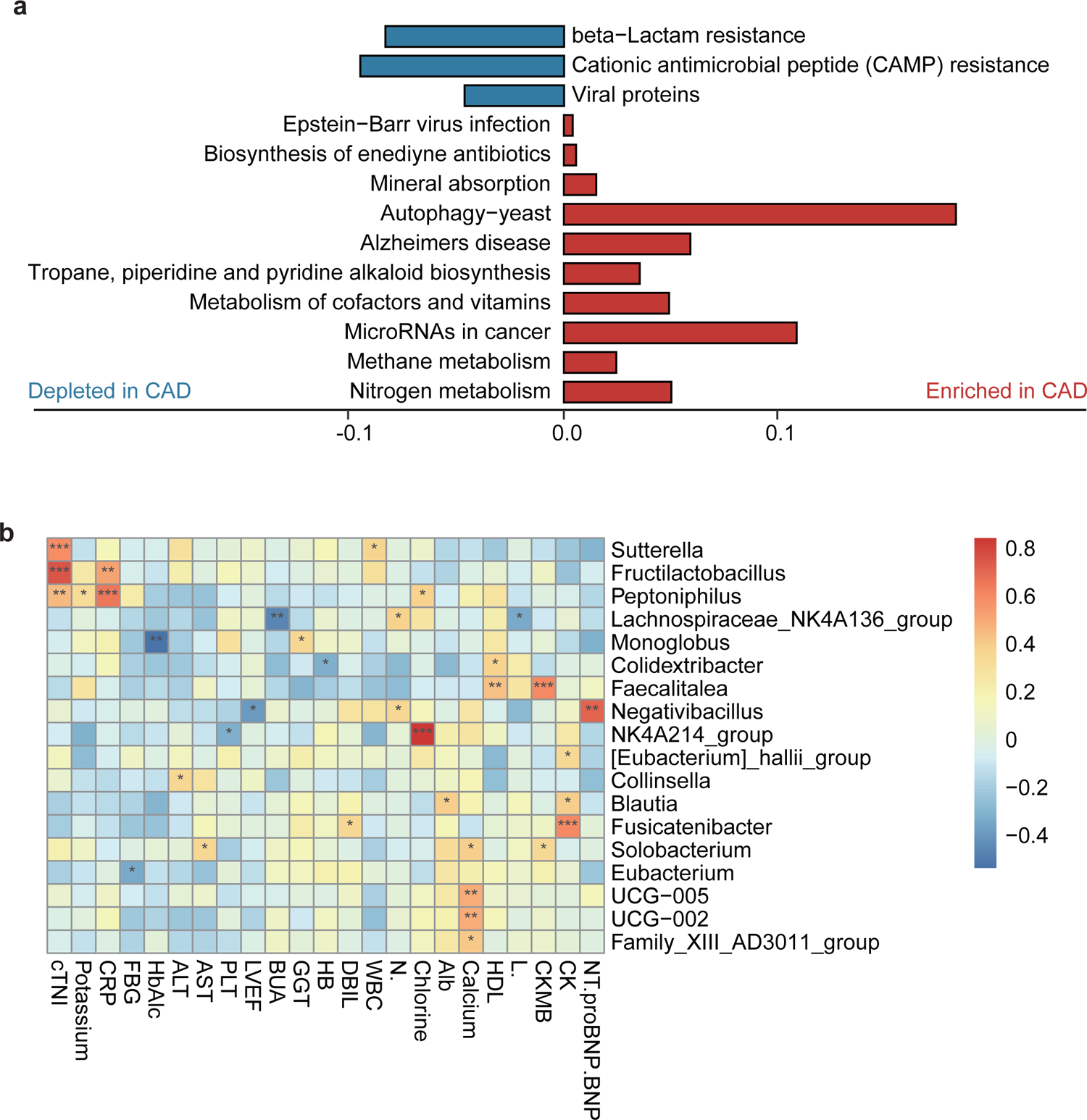
Function annotation and differential analyses of gut microbiota dysbiosis. (A) Functional implications of gut microbial dysfunction in patients with CAD; (B) The association of differential abundant bacteria with clinical indices.

Furthermore, we measured the associations of differential abundant bacteria between CAD and health groups with 23 clinical indices, including blood routine, heart, liver, and kidney functions. There were a series of significant associations between gut microbes and clinical indices relevant to heart functions. Genus *Sutterella*, *Fructilactobacillus*, and *Peptoniphilus* were positively associated with cardiac troponin I (cTNI), the cardiac regulatory proteins that control the calcium mediated interaction between actin and myosin. Brain natriuretic peptide (BNP) was significantly associated with genus *Negativibacillus*. Creatine kinase (CK) showed significantly positive correlations with genus *Fusicatenibacter*, *Blautia*, and *[Eubacterium]_hallii_group*. In addition, C-reactive protein (CRP) is a widely used marker of inflammation, and elevated CRP levels are involved in the development and progression of thrombosis and CAD. Our results indicate that CRP exhibit a significantly positive association with genus *Peptoniphilus* and *Fructilactobacillus* (Fig. 3b).

### Transcriptional analysis revealed genes associated with gut microbes

To reveal transcriptome difference between CAD patients and health controls, we further performed differential gene expression analysis on PBMC transcriptomic profiles. Comparing with health controls, there were 409 up-regulated and 762 down-regulated genes in CAD patients (P < 0.05; Fig. 4a), including *APOE* and *OLFML3,* which have been well-documented to be associated with CAD in previous studies [46, 47]. GSEA analysis showed that CAD group is characterized by depletion in interferon signaling-related pathways, including interferon gamma signaling pathway and interferon alpha/beta signaling pathway, and is enriched in several immune-regulated and inflammation-related pathways, including TNFs bind their physiological receptors, Rap1 signaling, and TNFR2 non-canonical NF-kB pathway (Fig. 4b).

**Fig 4.**
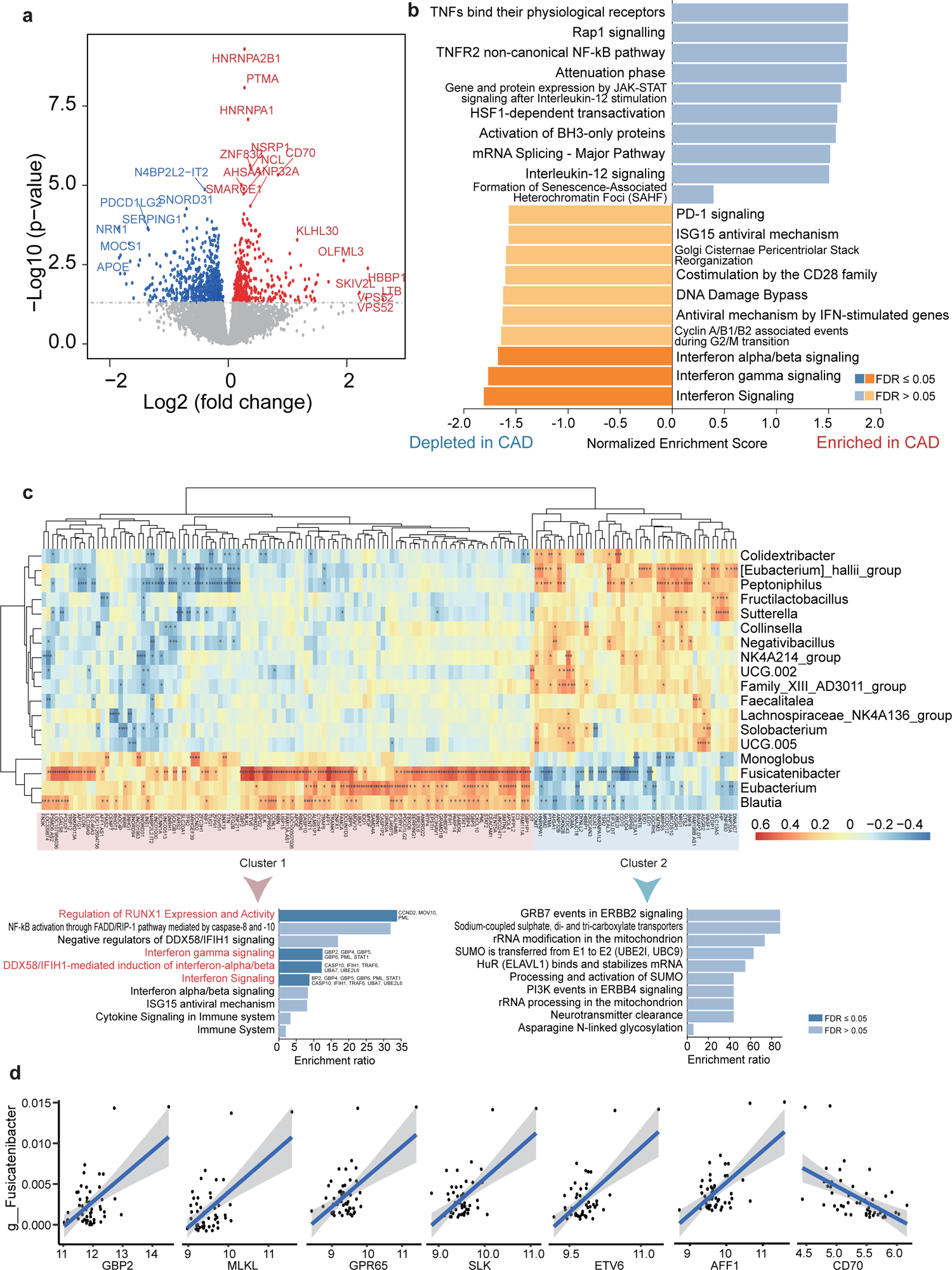
The transcriptome difference between CAD and control group. (A): The volcano map of differential gene; (B): The GSEA analysis of differential genes; (C) and (D): The correlation analysis on differentially abundant gut microbes and differentially expressed genes.

Considering that the significantly dis-regulated gut microbes might be associated with host gene abnormal expression and biological functions, we performed a correlation analysis on differentially abundant gut microbes (n=18) and differentially expressed genes (P < 0.01, n = 347). We obtained a total of 697 suggestive or significant correlations (P < 0.05, Fig. 4c). Among them, there were seven significant correlations with *g_Fusicatenibacter* with FDR < 0.05, including six positively correlated genes of *GBP2*, *MLKL*, *GPR65*, *SLK*, *ETV6*, and *AFF1*, and one negatively correlated gene of *CD70* (Fig. 4d). Among these genes, several have been documented to be implicated in the development of CAD [48, 49]. For example, a network-based prioritization analysis identified the interferon-induced guanylate-binding protein 2 (GBP2) as a key regulator orchestrating biological processes relevant to CAD [48]. *MLKL*, which can directly induce necroptosis [50], has been reported to be involved in different inflammatory diseases, including tumor necrosis factor-induced shock [51], ischemia-reperfusion injuries [52], and obesity [53]. Recently, Kamal and coworkers have found that *MLKL* is associated to hallmarks of atherosclerosis with and without type II diabetes mellitus, which could be a potential drug target for treating atherosclerotic patients [49]. Thus, these *g_Fusicatenibacter-*host gene interactions may play an important role in CAD.

### Different gene programs associated with gut microbes

We further performed differential gene program analysis to explore whether there exist distinct gene sets with different functions associated with gut microbes. Based on the unsupervised clustering analysis, these differential genes were grouped into two clusters (Fig. 4c). Cluster 1 (highlighted in red) were positively correlated with genera that were depleted in CAD patients (e.g., *Blautia*, *Eubacterium*, *Fusicatenibacter*, and *Monoglobus*). Cluster 2 (highlighted in blue) were positively correlated with genera that were enriched in CAD patients. Pathway enrichment analysis demonstrated that genes in cluster 1 showed significant enrichments in immune-related pathways (FDR < 0.05), including regulation of *RUNX1* expression and activity, interferon gamma signaling, *DDX58/IFIH1-*mediated induction of interferon-alpha/beta and interferon signaling. This highlighted a set of genes involved in interferon signaling pathway, such as *BP2*, *GBP4*, *GBP5*, *GBP6*, *PML*, *STAT1*, *CASP10*, *IFIH1*, *TRAF6*, *UBA7*, and *UBE2L6* (Fig. 4c). Although no significant enrichment was observed for genes in cluster 2 (FDR < 0.05), we identified that several pathways exhibited suggestive enrichments for genes in cluster 2 (P < 0.05), including *GRB7* events in *ERBB2* signaling, sodium-coupled sulphate, di- and tri-carboxylate transporters, and *PI3K* events in *ERBB4* signaling. Together, these two distinct gene programs relevant to gut microbes potentially have different biological functions contributing to CAD.

## Discussion

The role of gut microbes in CAD is still not fully understood. Deciphering correlation of gut microbes with CAD related gene functions and clinical indices contribute in understanding the underlying pathophysiological mechanism of CAD. In the current study, we performed 16S rRNA sequencing to comprehensively elucidate difference in profiles of gut microbiome in CAD patients and health controls. Our results exhibited that structure of gut microbiome differed in CAD and health group. In the CAD patients, 4 genera such as *Blautia*, *Eubacterium*, and *Fusicatenibacter* were depleted, and 14 genera such as *Sutterella*, and *Collinsella* were enriched.

Accumulating evidence has been documented that remarkably disturbed gut microbiota were detected in patients with cardiovascular diseases [28–31]. Mitra et al. [54] indicated that host microbiome associated families, such as *Porphyromonadaceae*, *Bacteroidaceae*, *Micrococcacaea*, and *Streptococcacaea*, were increased in asymptomatic atherosclerotic plaques patients than symptomatic atherosclerotic plaques patients. Based on these findings, we particularly focused on the relationship of altered *Blautia*, *Eubacterium*, and *Fusicatenibacter* with CAD. These three microbes were major producers of short chain fatty acids (SCFAs), which are important secondary metabolites capable of regulating cardiometabolic health [55–57]. Previous studies [58–60] have reported a reduced abundance of SCFA producers in CAD patients in different population and resolution. A recent study by Guo et al. [58] indicated that SCFA was significantly lower in AMI group than in control group. In addition, bacteria that can produce SCFA, such as *Ruminococcus* and *Bacteroides*, were significantly reduced in CAD patients [61].

Butyrate and other SCFAs produced by gut microbes serve as signaling molecules to modulate blood pressure, inflammatory responses and other metabolic functions. There are increasing evidence indicates the importance of SCFAs in regulating cardiac function [61, 62]. Considerable studies have demonstrated the parasympathetic activation effects of the SCFA propionate [61]. Zhou et al. [63] demonstrate that oral propionate supplementation improve MI therapy by parasympathetic activation based on the gut-brain axis. Moreover, a study of malonate intervention in the treatment of myocardial infarction in mice has showed that malonate enable to shift the cardiac metabolic pattern of oxidative phosphorylation to glucose metabolic pattern and promote cardiomyocyte proliferation, hemodialysis and cardiac regeneration in adult mice after myocardial infarction [64]. Jiang et al. [65] reported that intramembranous butyrate injection improved cardiac function by promoting macrophage differentiation and inhibiting inflammation and sympathetic remodeling after myocardial infarction. Our results emphasized the importance of gut microbial functions involved in cationic antimicrobial peptide (CAMP) resistance in CAD. CAMPs are hybrid peptides playing an important role in defensing against invasive bacterial infection for host. However, in addition to antimicrobial effect, CAMPs were believed to be an important link between inflammation and atherosclerotic cardiovascular disease [66]. For example, elevated plasma α-defensin is associated with an increased risk for cardiovascular morbidity [67]. PR-39 have been reported to play a cardioprotective role in myocardial I/R injury through inhibition of tumor necrosis factor-α (TNF-α)-induced degradation of the NFκB inhibitor IκBα [68].

To investigate the interactional functions of gut microbes and host genes on CAD, we performed correlation analysis of the abundances of gut microbes and the expression levels of host genes. We observed significant reduction of interferon signaling pathways in CAD patients, as well as strong association of depleted bacteria with genes involved in interferon signaling pathways. Our result suggested that depletion of SCFA-producing bacteria (*Blautia*, *Eubacterium*, and *Fusicatenibacter*) may contribute to the decrease of interferon signaling pathways in CAD patients. One mechanism by which microbes can modulate the host disease is through regulation of cytokine signaling [69]. Interferons (IFNs) are classified into three families (type I, type II, and type III) based on sequence homology. Among them, IFN-γ is a key cytokine implicated in both innate and adaptive immunity, and studies on the involvement of IFN-γ in multiple stages of the atherosclerotic process, playing different roles, have been carried out for decades. *In vitro* and *in vivo* studies [70–74] have shown that IFN-γ has both pro- and anti-atherogenic properties and plays a large role in all stages of CAD progression. Meanwhile, the type I interferons have the capacity to regulate the development or function of virtually every immune effector cell, contributing to the anti-inflammatory and anti-tumor responses, which is essential in the process of CAD [75].

## Conclusions

In summary, our results provide evidence to support that gut microbes play critical roles in CAD mediated by host genes. Reduction of SCFA-producing bacteria and interferon signaling genes are associated with the development of CAD. Depletion of SCFA-producing bacteria, e.g. *Blautia*, *Eubacterium*, and *Fusicatenibacter,* may contribute to dysfunction of interferon signaling and cardiovascular functions. These identified microbes may have potential for diagnosis or therapy of CAD.

## Methods

### Study participants and sample collection

In the current study, 31 patients with CAD and 23 healthy controls were recruited from the Department of Cardiology, Hangzhou First People’s Hospital, Zhejiang University School of Medicine. This study was reviewed and approved by the Ethics Committee of Zhejiang University. The enrolled patients signed a written informed consent form. All patients’ medical history and baseline data were obtained from the electronic medical records system. To obtain high-quality samples, patients with CAD included in this study were required to meet the following criteria: 1) age > 25 years and < 80 years; 2) show ≥70% stenosis in at least one major branch of the coronary artery. The healthy control group population should meet the following criteria: 1) age > 45 years and < 70 years; 2) all three coronary vessels ≤20% stenosis. Besides, individuals without coronary angiography testing or with one of follow disorders, including cancer, AMI, antibiotic exposure, and any other cardiac-related disease or systemic disease were excluded.

Pre-operative blood samples for these 54 participants were drawn and stored in PAXgene blood RNA tubes at −80°C before further processing [76]. During the hospitalization period, these subjects were educated and instructed on collecting fecal samples. All fresh fecal samples were stored in −80°C for subsequent processing and sequencing.

### 16S rRNA sequencing

Total genomic DNA was extracted from fecal samples. The V3-V4 region of bacterial 16S rRNA gene was amplified using primers (341F: 5′-CCTACGGGNGGCWGCAG-3′, 806R: 5′-GGACTACHVGGGTWTCTAAT-3′) with barcodes. The mixed PCR products were purified using the Qiagen Gel Extraction Kit (Qiagen, Germany). Sequencing libraries were generated using the TruSeq® DNA PCR-Free Sample Preparation Kit (Illumina, USA). The library quality was assessed using the Qubit@ 2.0 Fluorometer (Thermo Scientific) and the Agilent Bioanalyzer 2100 system. Finally, the library was sequenced on an Illumina NovaSeq 6000 platform, generating 250 bp paired-end reads.

Raw sequences were demultiplexed based on barcodes. After trimming off the barcodes and primers, paired-end sequences were assembled with FLASH (v1.2.7). The assembled raw tags were quality controlled with Fastp (v0.23.1) [77] to obtain high-quality clean tags. Chimeric sequences were further removed from clean tags using vsearch (v2.22.1) [78]. The filtered sequences were clustered based on 97% identity to generate Operational Taxonomic Units. The Uparse (v7.0.1001, http://www.drive5.com/uparse/) method was applied to cluster the filtered sequences into Operational Taxonomic Units (OTUs) at 97% identity threshold. Representative sequences were extracted and annotated with Mothur (v1.48.0) [79] and SILVA (v138.1) database [80]. The MUSCLE (v3.8.31) [81] tool was leveraged to perform sequence alignment for obtaining phylogenetic relationships of all OTU representative sequences. Finally, the data for each sample were normalized based on the sample with the lowest number of sequences, and subsequent alpha and beta diversity analyses were based on the normalized data.

### Transcriptomic sequencing

Total RNA was extracted from the isolated PBMCs using the QIAzol and miRNeasy Mini Kit (Qiagen, CA, USA). The RNA integrity was tested using the Bioanalyzer 2100 system with the RNA Nano 6000 Assay Kit (Agilent Technologies, CA, USA). Poly-T oligo-attached magnetic beads were used to purify mRNA from the total RNA, which was subsequently used to establish cDNA libraries for RNA sequencing. Quality-controlled cDNA library was sequenced on Illumina Novaseq platform (Beijing, China), producing paired-end sequences (150bp).

Raw transcriptomic sequencing data were qualified using the FastQC (https://www.bioinformatics.babraham.ac.uk/projects/fastqc/). Ensembl human reference genome (http://asia.ensembl.org/info/data/ftp/index.html, file name: Homo_sapiens.GRCh37.75.cdna.all.fa) was used for alignment and annotation. The Hisat2 (v2.0.5) tool [82] was used to establish index for reference genome, and align reads to the reference genome. The FeatureCounts (v1.5.0-p3) method [83] was used to summarize read counts for each gene.

### Functional enrichment analysis

We performed differential gene expression (DGE) analysis using the DESeq2 (v1.36.0) [84]. P-values were evaluated by the Student’s t-test [85], and multiple testing corrections were carried out using the Benjamini & Hochberg false discovery rate (FDR) method [86], analogue to a previous study [87]. Gene set enrichment analysis (GSEA) and over representative analysis (ORA) were performed by using the WEB-based Gene Set AnaLysis Toolkit (WebGestalt, https://www.webgestalt.org/) [88]. Pathways in the Reactome database [89] were used as a reference.

### Bioinformatics and Statistical analyses

Qiime (v1.9.1) [90] was used to calculate alpha diversity indexes and Unifrac distance and sample clustering tree. Unifrac distance measures the evolutionary branch weight of different OTUs in the sample OTU table to assess the differences between OTUs. After obtaining the Unifrac distance matrix, the UPGMA algorithm was used to construct a sample clustering tree. PCoA analyses was conducted with ade4 package in R (v2.15.3). Differences in alpha diversity indexes between groups were analyzed by the Wilcoxon rank-sum test [91] in R (v2.15.3).

Tax4Fun [92] was applied to predict functional profiles. Specifically, it extracted the full-length 16S rRNA gene sequences of prokaryotic genomes from the KEGG database [93–96] and used the BLASTN algorithm to align them to the SILVA SSU Ref NR database [97] (with a BLAST bitscore >1500) to establish a relevant matrix. The functional information of bacterial genomes from the KEGG database, annotated using the UProC and PAUDA methods, was then mapped to the SILVA database [80] for functional annotation. OTUs were clustered based on the SILVA database reference sequences.

Differences in bacterial composition and predicted functional profiles between groups were analyzed by the Student’s t-test [85] in R (v2.15.3). Additionally, we applied the Random Forest method [98] to recognize bacterial biomarkers that classify CAD patients from health controls. R package “randomForest” (RF) was used to perform random forest classification with default “mtry” parameters. Variable importance was measured by permuting each predictor variable and measuring the decrease in model accuracy. We adopted a nested cross-validation procedure [99] in which the least important variables were sequentially removed until the mean error rate of the model reached two standard deviations above the lowest error rate. We calculated the area under the ROC curve (AUC) with the “ROCR” package [100] in R (v2.15.3). To select biomarkers, we sorted gut microbes according to their RF importance and calculated the AUC using different numbers of top gut microbes. The gur microbes with the highest AUC were selected as biomarkers.

To assess the correlations of bacterial abundances with level blood indicators and expression levels of genes in paired peripheral blood, the Pearson correlation test was applied, and FDR < 0.05 was considered to be significant. All the plots were visualized in R (v2.15.3).

## Abbreviations

CAD: coronary artery disease

TMAO: trimethylamine-N-oxide

CAMP: cationic antimicrobial peptide

BNP: brain natriuretic peptide

cTNI: cardiac troponin I

CK: creatine kinase

CRP: C-reactive protein

SCFA: short chain fatty acids

PBMCs: peripheral blood mononuclear cells

DGE: differential gene expression

FDR: false discovery rate

GSEA: gene set enrichment analysis

ORA: over representative analysis

KEGG: the Kyoto Encyclopedia of Genes and Genomes

AUC: area under the ROC curve.

## Data Availability

The original contributions presented in the study are included in the article/Supplementary Material. Detailed clinical baseline data is available at BIG Submission (BIG SUB, https://ngdc.cncb.ac.cn/omix/preview/FQcgNvTM, OMIX ID:OMIX004322).

## Acknowledgements

We thank all participants and investigators who provide a contribution to the work.

## Authors’ contributions

CL, MX, and LJ conducted the experiments and wrote the article. LM, YC, GX, and LX were involved in parts of the experiments. XY, MY, and ZY conceived the hypothesis, designed the experiments, and reviewed the article. ZY and MY helped in guiding the revision. All authors reviewed and approved the final article.

## Funding

This research was funded by the Hangzhou Medical and Health Technology Project (No. Z20200135), the Construction Fund of Key Medical Disciplines of Hangzhou (No. OO20200121), the National Natural Science Foundation of China (No. 32200535), and the Scientific Research Foundation for Talents of Wenzhou Medical University (No. KYQD20201001).

## Declarations

### Ethics approval and consent to participate

All procedures followed were in accordance with the ethical standards of the responsible committee on human experimentation (institutional and national) and with the Helsinki Declaration of 1975 (in its most recently amended version). Informed consent was obtained from all patients included in the study.

### Consent for publication

Not applicable

### Competing interests

The authors declare no competing interests.

## Notes

### Competing Interest Statement

The authors have declared no competing interest.

### Author Declarations

This study was reviewed and approved by the Ethics Committee of Zhejiang University. The enrolled patients signed a written informed consent form.

