## Supplemental Figure for "Alternations in gut microbiota and host transcriptome of patients with coronary artery disease"

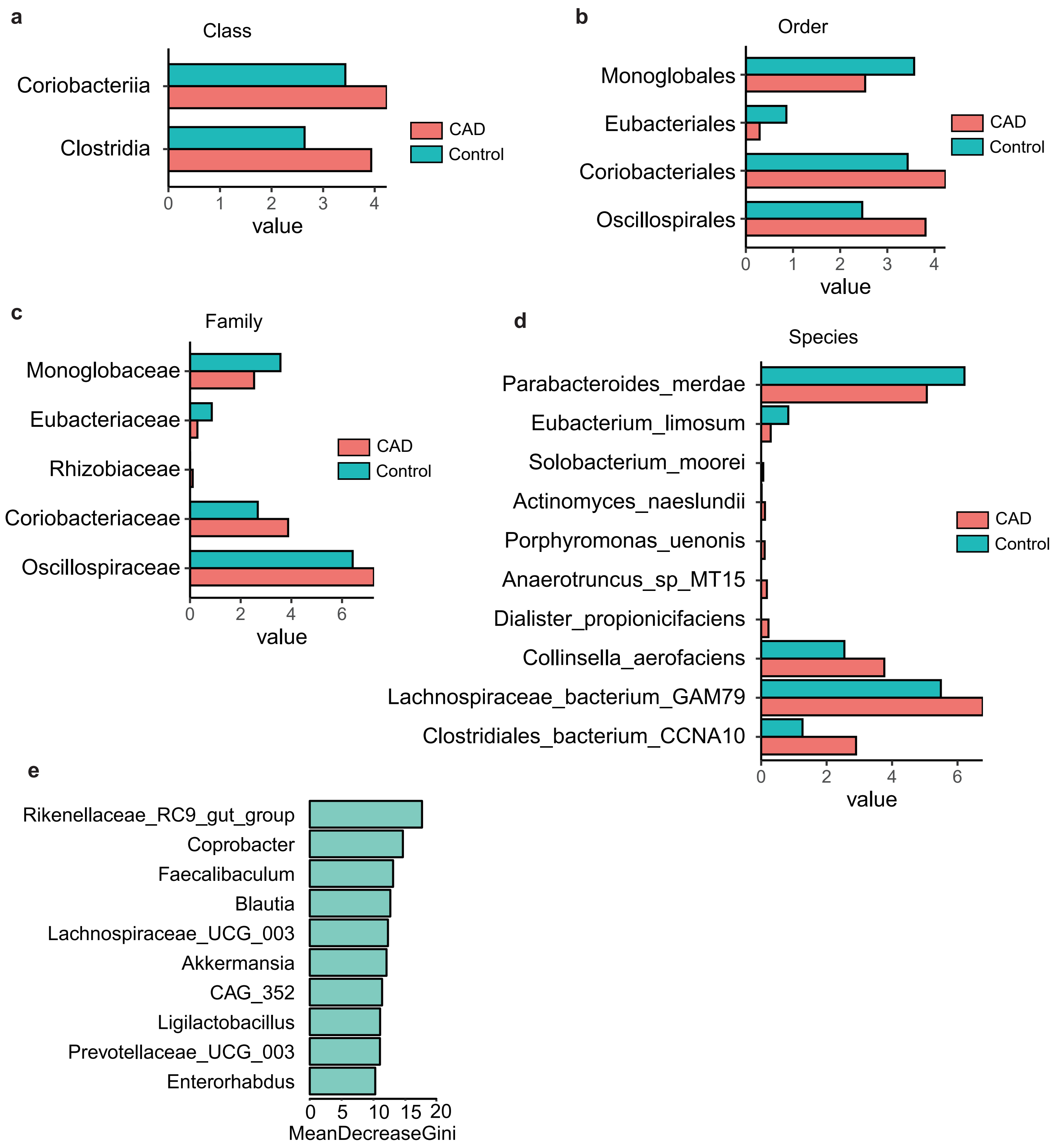


**Supplementary Figure S1**. Difference of gut microbiota between CAD and Control group at class (A), order (B), family (C), and species (D) level. (E) The top 10 differential genera between CAD and control group by means of Mean Decrease Gini.
